# Parafoveal Dark Adaptation in Early and Intermediate Age-Related Macular Degeneration

**DOI:** 10.1101/2025.11.21.25340690

**Authors:** Georg Ansari, Jeannine Oertli, Laura Mächler, Theresa Lipsky, Brett G Jeffrey, Catherine A Cukras, Nicolas Feltgen, Caroline C W Klaver, Kristina Pfau, Maximilian Pfau

**Affiliations:** Department of Ophthalmology, University Hospital Basel, Switzerland; National Eye Institute, National Institutes of Health, Bethesda, MD, USA; Institute of Molecular and Clinical Ophthalmology Basel (IOB), Switzerland; Department of Ophthalmology, Erasmus University Medical Center, Rotterdam, The Netherlands; Department of Ophthalmology, University Hospital Bonn, Germany

**Keywords:** Fundus-controlled dark adaptometry, dark adaptation, early AMD, intermediate AMD, biomarker, functional endpoints

## Abstract

**Purpose:** Rod-mediated dark adaptation delays are among the earliest functional abnormalities in age-related macular degeneration (AMD), preceding photoreceptor loss. This study evaluated whether parafoveal fundus-tracked dark adaptometry detects earlier rod dysfunction than more eccentric mid-macula testing and assessed the diagnostic performance of dynamic and steady-state parameters across eccentricities.

**Methods:** In this cross-sectional study, 35 patients with predominantly early / intermediate AMD and 32 age-spanning controls underwent fundus-controlled dark adaptometry (S-MAIA-2; iCare/CenterVue) and multimodal imaging. After standardized bleaching, cyan stimuli were presented 2°, 4°, and 6° temporal to the fovea. Dark-adaptation curves were modeled to derive rod intercept time (RIT), final rod threshold (FT), and cone threshold (CT), each compared with age-adjusted normative data. Diagnostic accuracy was quantified using age-adjusted receiver operating characteristic (ROC) analyses.

**Results:** Among 67 analyzed eyes, RIT was abnormal in 89% of AMD eyes at 2°, 77% at 4°, and 74% at 6°, whereas FT and CT were less frequently abnormal (29 to 51% and 17 to 23%, respectively). Median RIT at 2° reached 60 minutes, indicating incomplete recovery in many eyes. RIT achieved the highest diagnostic accuracy, with age-adjusted AUC values of 0.91 (95% CrI, 0.81–0.98) at 2°, 0.88 (0.77–0.96) at 4°, and 0.87 (0.76–0.95) at 6°.

**Conclusions:** Fundus-tracked dark adaptometry enables spatially precise assessment of parafoveal rod recovery. Parafoveal RIT prolongation represents the earliest and most frequent functional abnormality in AMD and demonstrates excellent diagnostic performance, supporting its potential as a sensitive functional biomarker for early disease and therapeutic trials.

## INTRODUCTION

Age-related macular degeneration (AMD) is the leading cause of legal blindness in developed countries, with limited therapeutic options currently available.^1,2^ Globally, the prevalence of any form of AMD has been estimated at 196 million, projected to rise to 288 million in 2040.^2^

AMD initially affects Bruch’s membrane and the choriocapillaris, resulting in a characteristic appearance resembling an ‘oil spill’ in histopathology.^3,4^ With time, these deposits coalesce to form drusen within the inner layers of Bruch’s membrane.^3,4^ Enlargement of such drusen together with pigmentary abnormalities indicate an increased likelihood of progression toward late-stage disease.^5–10^ Late-stage disease is characterized by two major trajectories: either geographic atrophy, marked by the loss of retinal pigment epithelium and overlying photoreceptors, or neovascular AMD, characterized by angiogenesis and exudation, both of which culminate in central vision loss.

Microstructural investigations – histopathology and in vivo imaging studies – converge on a parafoveal annulus, extending between eccentricities of approximately 750 µm (2.6°) and 1,250 µm (4.3°), as the region of greatest susceptibility for the incidence of geographic atrophy.^11,12^ This ring also corresponds closely to the broader parafoveal ring spanning 0.5 to 3 mm eccentricity, where rod photoreceptor density undergoes its steepest decline with aging and AMD.^13,14^

Psychophysical studies likewise localize early rod dysfunction to the parafovea, where rod-mediated vision is disproportionately vulnerable to aging and AMD.^12,15–19^ As early as 1993, Steinmetz and coworkers reported delayed rod-mediated dark adaptation in AMD patients, i.e., *dynamic* dysfunction of the rod system occurring even when *steady-state* thresholds remained normal.^18^ Further, a gradient of increasing impairment was observed toward the fovea (closest test locus at 3° eccentricity).^18^ Subsequent investigations confirmed this pattern using multifocal dark adaptometry (testing as close as 4° eccentricity),^20,21^ or repeated unifocal dark adaptometry (comparing 5° and 12°).^22^ However, quantitative data characterizing dark-adaptation delays in the actual region of interest – the parafoveal ‘high-risk’ annulus – remain absent to date.

Recently, the advent of fundus-tracked dark adaptometry has enabled localized assessment of rod-mediated vision within the parafovea. Active fundus tracking reduces reliance on precise fixation by participants, allowing the use of dim fixation lights that minimize interference with dark adaptation. Furthermore, using small stimuli facilitates spatially specific testing (i.e., Goldmann III [0.43° diameter] stimuli instead of Goldmann V [1.72°] or 2° stimuli). Together, these advances provide the capability to probe rod function specifically within the parafoveal annulus, identified microstructurally as the earliest region of susceptibility in AMD.

As the logical next step, we evaluated the feasibility of fundus-tracked dark adaptometry in AMD, compared outcomes to age-spanning controls, and assessed its diagnostic validity across dark-adaptation parameters and stimulus eccentricities. To establish concurrent validity, we performed structure–function analyses with established structural biomarkers of AMD severity. Finally, we examined the inter-relationships among dark-adaptation parameters to explore whether the non-linear associations of functional loss are indicative of a functional disease sequence.

## METHODS

### Study Design

In this prospective study, healthy volunteers and patients with AMD were recruited and examined in a tertiary referral center (University Hospital Basel). The study was approved by the ethics committee (BASEC ID: 2022-01243 [https://raps.swissethics.ch/]) and conducted in accordance with the Declaration of Helsinki. All participants were informed of the study’s nature and provided written informed consent before participating in study-related examinations.

Participants included eyes with early or intermediate AMD (per eye Beckman classification) and a small subset with geographic atrophy (GA).^23^ GA eyes were required to have an intact retina at the test locations (2°, 4°, and 6° temporal to the fovea) to allow meaningful measurement of dark adaptation. GA eyes were included to extend the functional spectrum of disease severity.

Other inclusion criteria for AMD patients encompassed visual acuity of <1.0 logMAR, clear optic media, and no treatment with anti-VEGF in study eye. Exclusion criteria were the inability to provide informed consent, severe claustrophobia, previous ocular surgeries (other than cataract surgery, YAG laser capsulotomy, refractive laser surgery), and concurrent ophthalmic conditions in the study eye that might independently affect visual acuity, as judged by the investigator.

The study eye was selected according to a predefined procedure. Eyes with no prior history of exudative macular neovascularization (i.e., without previous anti-VEGF treatment) were chosen. If both eyes met this criterion, the eye with the superior BCVA was designated as the study eye. In cases where BCVA was identical in both eyes, a random eye was selected.

### Imaging: Spectral-Domain Optical Coherence Tomography (SD-OCT), Fundus-Autofluorescence and Color Fundus Photography

SD-OCT imaging of the macula was obtained with a Heidelberg Spectralis device (Heidelberg Engineering, Heidelberg, Germany) with a 30° × 25° (121 B-scans, HR mode, enhanced Automatic Real Time-Function (ART) averaged 25 scans). Short-wavelength fundus autofluorescence (FAF) images (30° × 30°, HS mode, ART 20) were performed using the same device. True-color color fundus photography (CFP) was obtained with the Zeiss Clarus 500 device (Carl Zeiss AG, Jena, Germany).

### Fundus-controlled Dark-adaptometry

Patients underwent multifocal fundus-controlled dark adaptometry using the S-MAIA-2 device (iCare/CenterVue, Padova, Italy). The protocol used a full-field bleach (634 photopic cd/m², equivalent to 946 scotopic cd/m²) lasting 5 minutes (equivalent to a 59% rhodopsin bleaching, or a 54% cone pigment bleach, see appendix in Flynn et al. for calculation)^21^ from the MonCvONE device (Metrovision, Perenchies, France). Afterwards, patients were quickly moved to the S-MAIA-2 device. Cyan and red stimuli (peak wavelengths of 500 nm and 647 nm, stimulus duration 200 ms) were presented at retinal loci 2°, 4°, and 6° temporal to fixation (Figure 1). Patient were tested continuously in a 5 up 1 down strategy, i.e. the stimulus was presented 5 decibels (dB) darker after a response and 1 dB brighter if no response was recorded. If subjects reported fatigue during the test, short breaks were made for one minute during the tests to avoid prolonged recording breaks. Testing continued for up to 60 minutes or was terminated earlier if steady-state rod thresholds were achieved at all loci.

**Figure 1.**
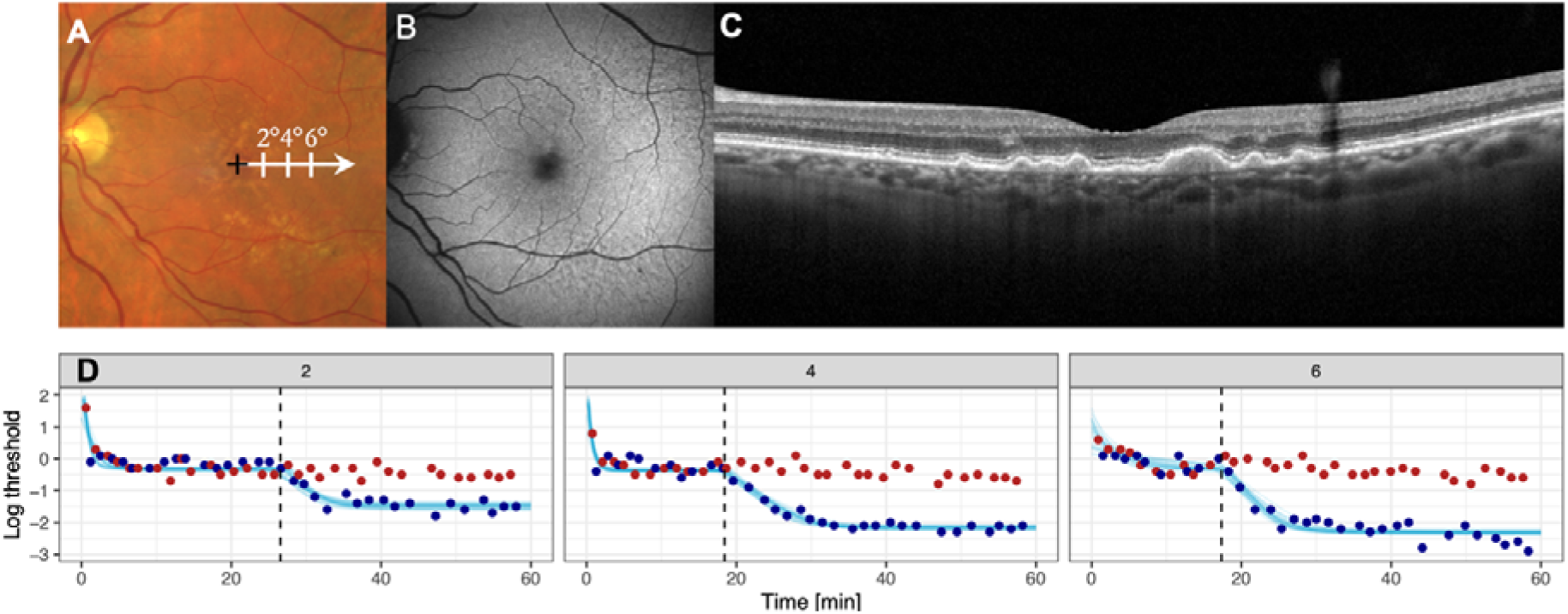
Fundus-controlled Dark Adaptometry in Age-related Macular Degeneration (AMD). The panels A to C show multimodal imaging of a representative patient with color fundus photography (A), fundus autofluorescence (B), and spectral-domain optical coherence tomography (C) imaging. Panel A illustrates the stimulus locations along the horizontal meridian at 2°, 4°, and 6° temporal to the fovea (indicated by the black cross). Panel D displays the corresponding dark-adaptation curves for red (red dots) and cyan (blue dots) stimuli at eccentricities of 2°, 4°, and 6 °. The y-axis indicates retinal sensitivity (log threshold), and the x-axis shows time (minutes) after the initial bleaching. Sky-blue lines represent fitted model curves; the vertical dashed line denotes the cone–rod break. The foveal location was estimated visually in the color fundus photography for illustrative purposes only and was not involved in any of the quantitative analyses.

Dark adaptation curve estimations were modeled using a Bayesian nonlinear regression with the R package *brms*.^24^ The curves were fitted using a biphasic model defined as follows:

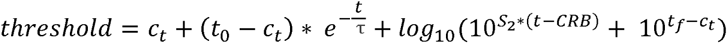

if time > CRB, otherwise:

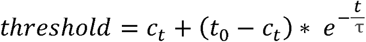

with the parameters: c_t_ = cone threshold, t_O_= initial threshold at time 0, τ = exponential time constant, S_2_ = rod adaptation slope, CRB = cone-rod break, and t_f_ = final (rod) threshold, all estimated as nonlinear terms (Supplementary Figure S1). We applied weakly informative priors for the fitting that spanned the plausible value range for the curve parameters (Supplementary Table S1).

Using nonlinear curve fitting, we determined cone thresholds (CT) and final (rod) thresholds (FT) in logUnits. The S-MAIA-2 device presents stimuli across a dynamic range of 0 to 36 dB, where a 10 dB change correspond to 1 logUnit change in luminance (cd/m^2^). The maximum stimulus intensity is defined as 0 dB, corresponding to 318 cd/m^2^.^25^ Rod intercept time (RIT) was measured in minutes. We chose −1.4 logUnits as the criterion threshold for RIT because it lies approximately 1 logUnit below the normal cone threshold. The researcher manually reviewed all curves. If there was no evidence of a cone-rod break (i.e. no separation between cyan and red stimuli) within the 60-minute test, the final threshold was considered cone-mediated, and RIT was recorded as 60 minutes.

### Statistical Analysis

Statistical analyses were performed in *R* using the add-on packages *tidyverse*,^26^ *ggplot2*,^27^ *table1*,^28^ *tidymodels*,^29^ *dplyr*,^30^ and *caret*.^31^

To estimate age-adjusted normal limits from the healthy volunteer data, we applied Bayesian linear mixed-effects regression models to estimate the relationship between a given dependent variable (cone threshold, final threshold, RIT) and age and retinal eccentricity. Age was modeled as a fixed effect, retinal position as a fixed categorical effect, and subject-specific variability was accounted for by including a random intercept for each participant. Weak priors were specified based on previously published normative datasets.^32,33^ The predicted values were summarized by a median and 95% prediction interval. To evaluate the diagnostic accuracy of cone threshold, final threshold, and RIT for distinguishing between healthy individuals and AMD patients at different eccentricities, covariate-adjusted receiver operating characteristic (ROC) curves with age as a covariate were generated using the R package *ROCnReg.*^34^ Adjustment for age ensured that the diagnostic performance estimates were not confounded by the disparity in age range between groups (i.e., yielding a more conservative estimate than ‘pooled’ ROC analyses). To explore interrelationships among dark-adaptation parameters, the parameters were expressed relative to age-adjusted normative limits: i.e., RIT in terms of RIT delay and cone threshold, and final threshold in terms of threshold deviation. Scatter plots were generated to visualize nonlinear dependencies and to identify potential thresholds beyond which steady-state sensitivity declined.

For structure-function analysis, we evaluated the relationship between RIT and established biomarkers of AMD severity (AMD severity of the study eye and fellow eye [early, intermediate, late - according to the Beckman classification], presence/absence of SDD in the study eye, and age) using linear regression analysis.

## RESULTS

### Cohort Characteristics

Thirty-five patients with clinically confirmed AMD in both eyes, with a median [IQR] age of 70.9 years [65.5, 79.3], and 32 healthy volunteers (50.6 [27.1, 62.6] years) were included in this study (Table 1). Median visual acuity in the study eye was −0.07 logMAR [−0.07, 0.11] in the AMD cohort and −0.07 logMAR [−0.07, −0.07] in healthy individuals. Subretinal drusenoid deposits (SDD) were present in 10 (28.6%) AMD patients (Supplementary Table S2, Supplementary Table S3).

**Table 1.** Cohort Characteristics.

|  | AMD<br>(n=35) | Healthy volunteers<br>(n=32) |
| --- | --- | --- |
| **Age [years]** |  |  |
| Median [IQR] | 70.9 [65.5, 79.3] | 50.6 [27.1, 62.6] |
| **Sex** |  |  |
| Female | 22 (62.9%) | 19 (59.4%) |
| Male | 13 (37.1%) | 13 (40.6%) |
| **Study eye** |  |  |
| Left | 14 (40.0%) | 20 (62.5%) |
| Right | 21 (60.0%) | 12 (37.5%) |
| **Subretinal drusenoid deposits** |  |  |
| Absent | 25 (71.4%) | NA |
| Present | 10 (28.6%) | NA |
| **Study eye visual acuity [logMAR]** |  |  |
| Median [IQR] | -0.07 [-0.07, 0.11] | -0.07 [-0.07, -0.07] |
| **Diagnosis: Study eye** |  |  |
| eAMD | 11 (31.4%) | 0 (0%) |
| iAMD | 20 (57.1%) | 0 (0%) |
| late AMD: GA | 4 (11.4%) | 0 (0%) |
| None | 0 (0%) | 32 (100%) |
| **Diagnosis: Fellow eye** |  |  |
| eAMD | 10 (28.6%) | 0 (0%) |
| iAMD | 17 (48.6%) | 0 (0%) |
| late AMD: GA | 6 (17.1%) | 0 (0%) |
| late AMD: nAMD | 2 (5.7%) | 0 (0%) |
| None | 0 (0%) | 32 (100%) |

*eAMD = early AMD, iAMD = intermediate AMD, nAMD = neovascular AMD, GA = geographic atrophy*

### Dark Adaptation in the Parafovea versus Mid-Macula

Cone-mediated thresholds were largely preserved across eccentricities. Median [IQR] steady-state cone thresholds were −0.49 logUnits [−0.63, −0.32] in the AMD cohort, corresponding to a median cone threshold deviation from age-adjusted normative predictions of −0.07 logUnits [−0.23, 0.05] at 2°, −0.04 logUnits [−0.24, 0.06] at 4°, and −0.07 logUnits [−0.29, 0.07] at 6°. Considering the 95% prediction intervals derived from healthy volunteers, cone thresholds fell outside the normal limits in only a minority of AMD eyes: 6 (17%) at 2°, 8 (23%) at 4°, and 9 (26%) at 6° (Figure 2).

**Figure 2.**
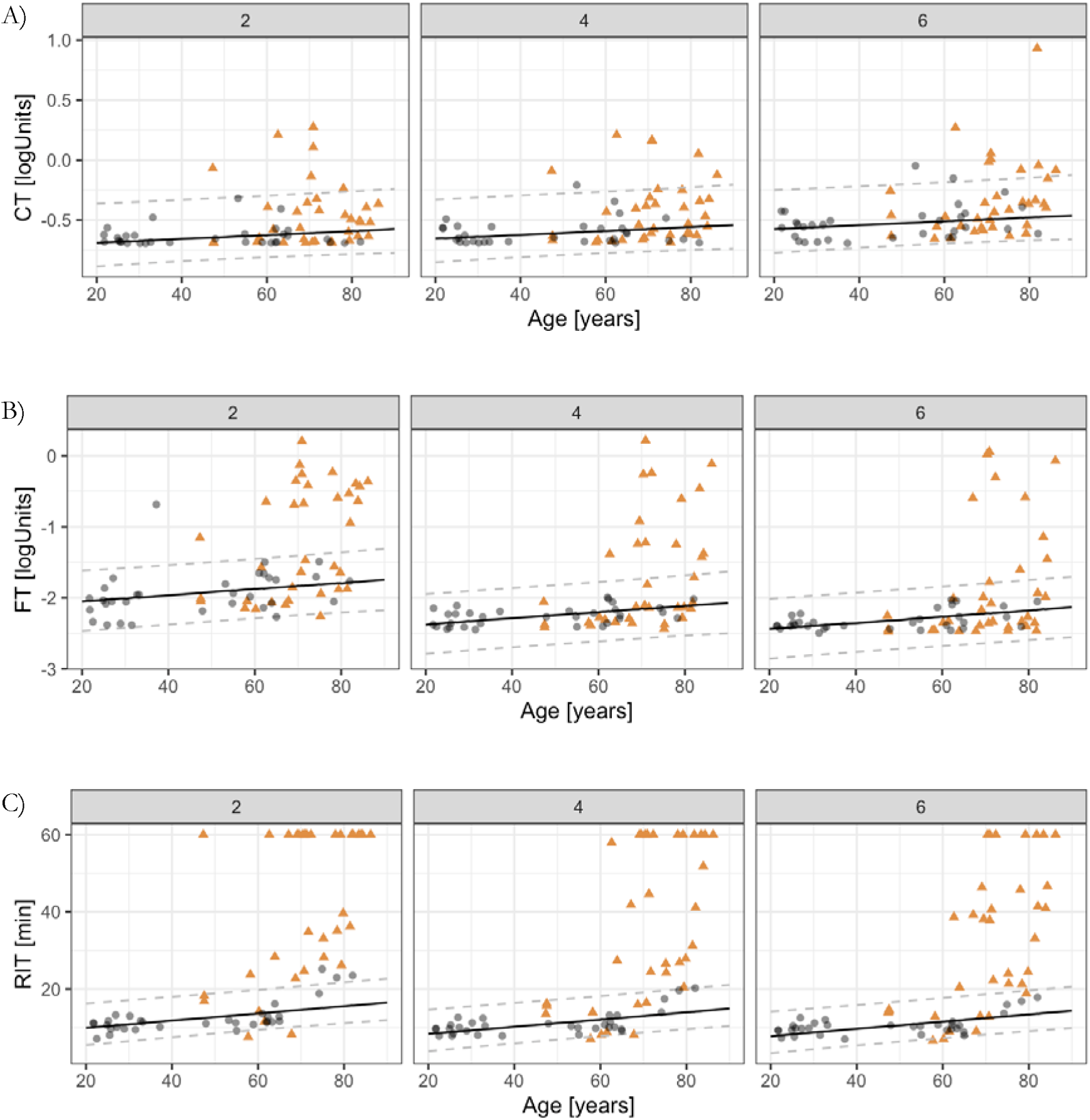
Dark Adaptometry Outcomes in Age-Related Macular Degeneration (AMD) and Healthy Controls. Cone threshold (CT), final (rod) threshold (FT), and rod intercept time (RIT) are plotted against age for test loci at 2°, 4°, and 6° eccentricity. Gray circles represent healthy controls, and orange triangles represent patients with AMD. Solid lines indicate the estimated mean random-slope model for healthy participants, with dashed lines showing the corresponding 95% prediction intervals.

Rod mediated steady-state thresholds were −1.99 logUnits [IQR −2.29, −0.69] in AMD patients, corresponding to a median rod threshold deviation from age-adjusted normative predictions of −0.35 logUnits [−1.29, 0.10] at 2°, −0.04 logUnits [−0.90, 0.12] at 4°, and 0.07 logUnits [−0.51, 0.17] at 6°. In contrast to cone thresholds, rod-mediated steady-state thresholds were more frequently abnormal, exceeding age-adjusted limits in 18 (51%) eyes at 2°, 13 (37%) at 4°, and 10 (29%) at 6° temporal to the fovea.

The most striking dysfunction, however, emerged in the kinetics of rod recovery. RITs were markedly prolonged in AMD, most severe at 2° eccentricity, with a median [IQR] of 60 minutes [25.4, 60.0], indicating incomplete recovery in many eyes. Recovery improved toward the mid-macula, with medians of 31.2 minutes [16.3, 60.0] at 4° and 33.1 minutes [14.4, 46.0] at 6°.

When compared to age-adjusted normative predictions, RIT was prolonged by a median [IQR] of 43.98 minutes [10.49, 45.33] at 2°, 17.20 minutes [5.05, 45.88] at 4°, and 19.64 minutes [4.13, 32.71] at 6°. RIT fell outside normal limits in 31 (89%) eyes at 2°, 27 (77%) at 4°, and 26 (74%) at 6° (Supplementary Table S4).

### Discriminating Accuracy between AMD and Healthy Controls

Receiver operating characteristic analyses adjusted for age demonstrated that RIT most effectively distinguished eyes with AMD from healthy controls at all tested eccentricities (Figure 3). Discriminative performance was highest at 2°, with an age-adjusted ROC area under the curve (ROC-AUC) of 0.91 (95% CrI, 0.81, 0.98), and remained robust at 4° (0.88 [0.77, 0.96]) and 6° (0.87 [0.76, 0.95]).

**Figure 3.**
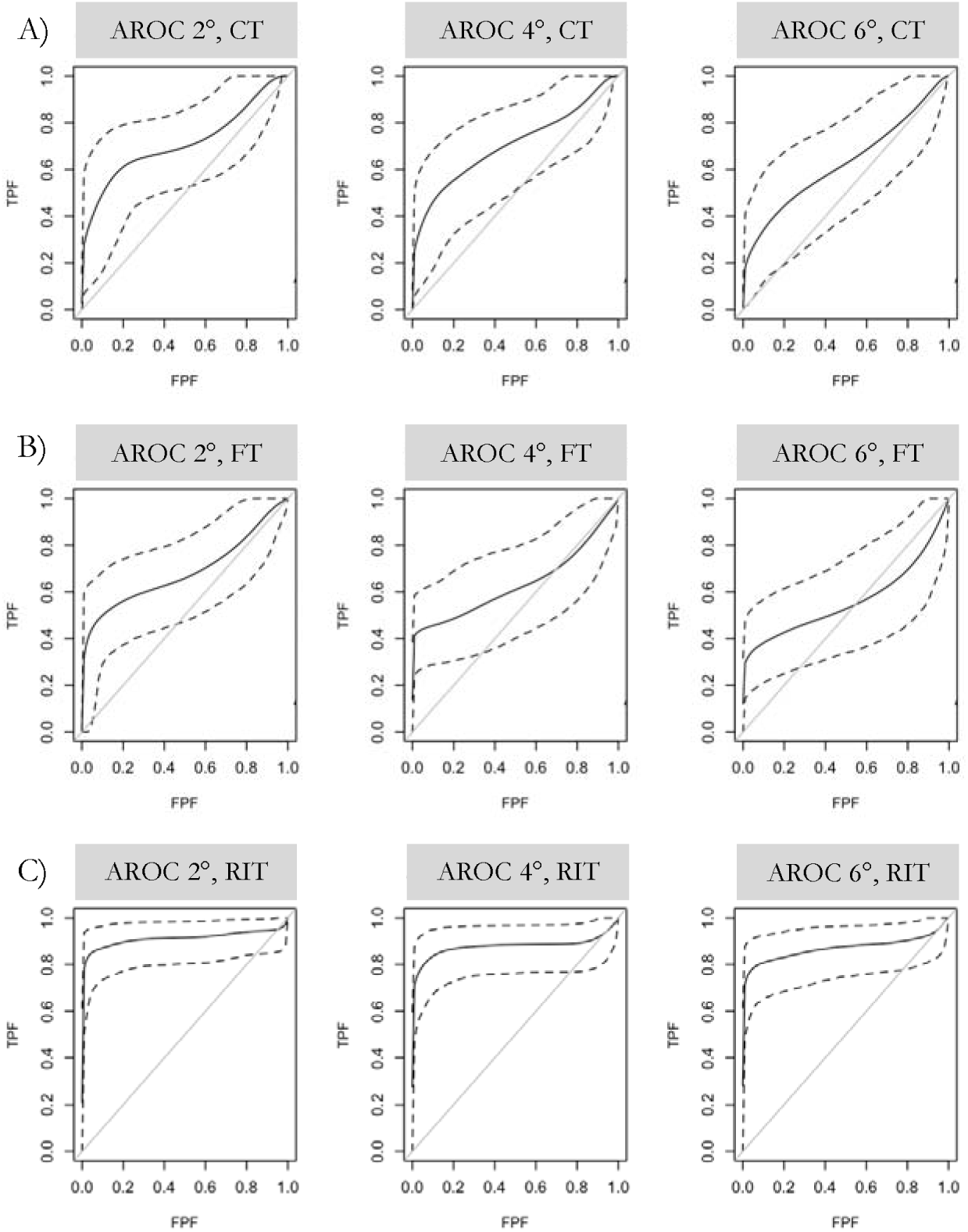
Age-Adjusted Receiver Operating Characteristic (ROC) Curves for Differentiating AMD from Healthy Controls. ROC curves are shown for cone threshold (CT), final rod threshold (FT), and rod intercept time (RIT) (rows) across test eccentricities of 2°, 4°, and 6° (columns). Each panel plots the true positive fraction (TPF) against the false positive fraction (FPF). Solid lines represent mean ROC curves, and dashed lines indicate the corresponding 95% credible intervals.

At every test location, the dynamic measure of rod recovery (RIT) surpassed steady-state thresholds in separating disease from normal. Final rod threshold yielded lower ROC-AUCs (0.69 [0.54, 0.82], 0.63 [0.48, 0.78], and 0.56 [0.41, 0.72]), and cone thresholds performed only modestly as well, 0.71 (0.57, 0.84), 0.71 (0.55, 0.83), and 0.63 (0.46, 0.78).

### Interrelationships Among Dark Adaptation Parameters

The relationships between dynamic and steady-state measures of dark adaptation were nonlinear (Figure 4A and B). When RIT delays were modest (<30 minutes), final (rod) thresholds were generally preserved, with final threshold deviations clustering near normal limits. Except for two outlier loci, no substantial final threshold deviations occurred within this range. Beyond an RIT delay of approximately 40 minutes, however, nearly all eyes exhibited marked deviations of the final threshold, compatible with an inflection point beyond which rod recovery failure transitions into measurable steady-state dysfunction.

**Figure 4.**
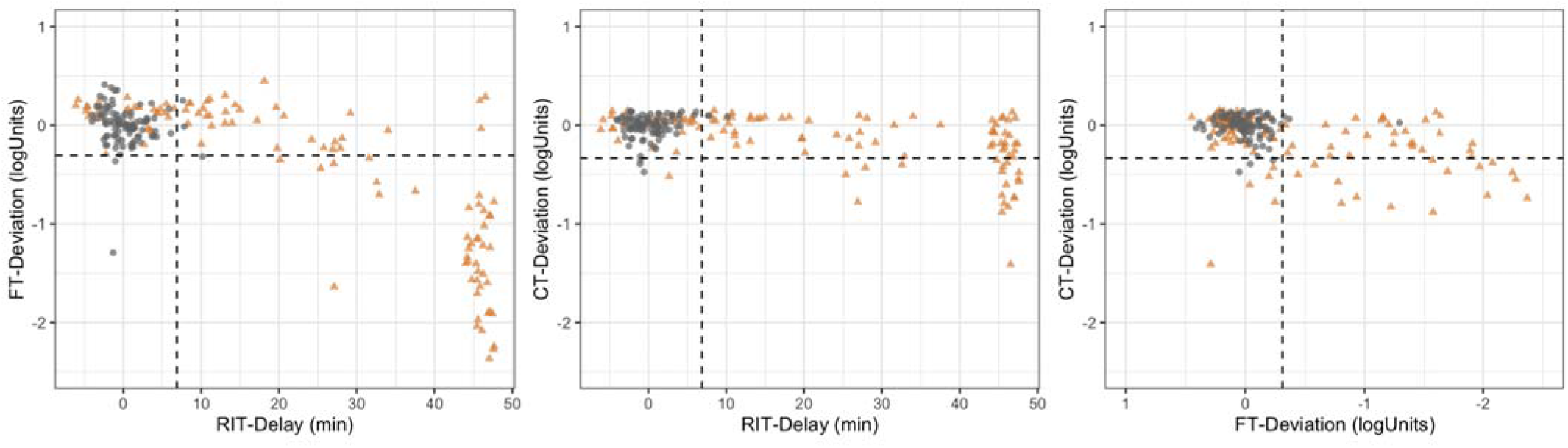
Pairwise Associations between Dark Adaptometry Outcomes in Patients with Age-related Macular Degeneration and Healthy Controls. The left panel shows the relationship between rod-intercept time (RIT)-delay and final rod threshold (FT)-deviation. The middle panel illustrates the association between RIT-delay and cone threshold (CT)-deviation. The right panel depicts the relationship between FT-deviation and CT-deviation. The grey circles represent healthy controls, while orange triangles represent patients with AMD. Dashed lines indicate the estimated 97.5% percentile (upper quantile) of the healthy cohort for RIT-delay, FT-deviation and CT-deviation.

A similar but less pronounced trend was observed for cone threshold. Cone threshold deviations were observable at locations with RIT delays of 40 minutes or longer, but with a smaller magnitude compared to the final (rod) threshold deviations. The relationship between cone threshold and final (rod) threshold deviations was linear, implying that steady-state deficits occur concurrently.

### Structure-Function Associations

Multivariate regression (Table 2) identified subretinal drusenoid deposits as the strongest determinant of delayed rod recovery, with RIT prolongations of +18.8 minutes at 2°, +24.4 minutes at 4°, and +26.4 minutes at 6° (all p < 0.001). Intermediate and late AMD were independently associated with additional RIT delays of 21–34 minutes across all eccentricities (all p < 0.001), whereas early AMD exerted no significant effect. Age modestly increased RIT (1.56–2.07 minutes per decade; p = 0.03 – 0.04). Fellow-eye diagnosis showed no consistent association with RIT across eccentricities. The models accounted for 71%–79% of total RIT variance, underscoring the strong, coherent relationship between retinal structure and rod-mediated function.

**Table 2.**
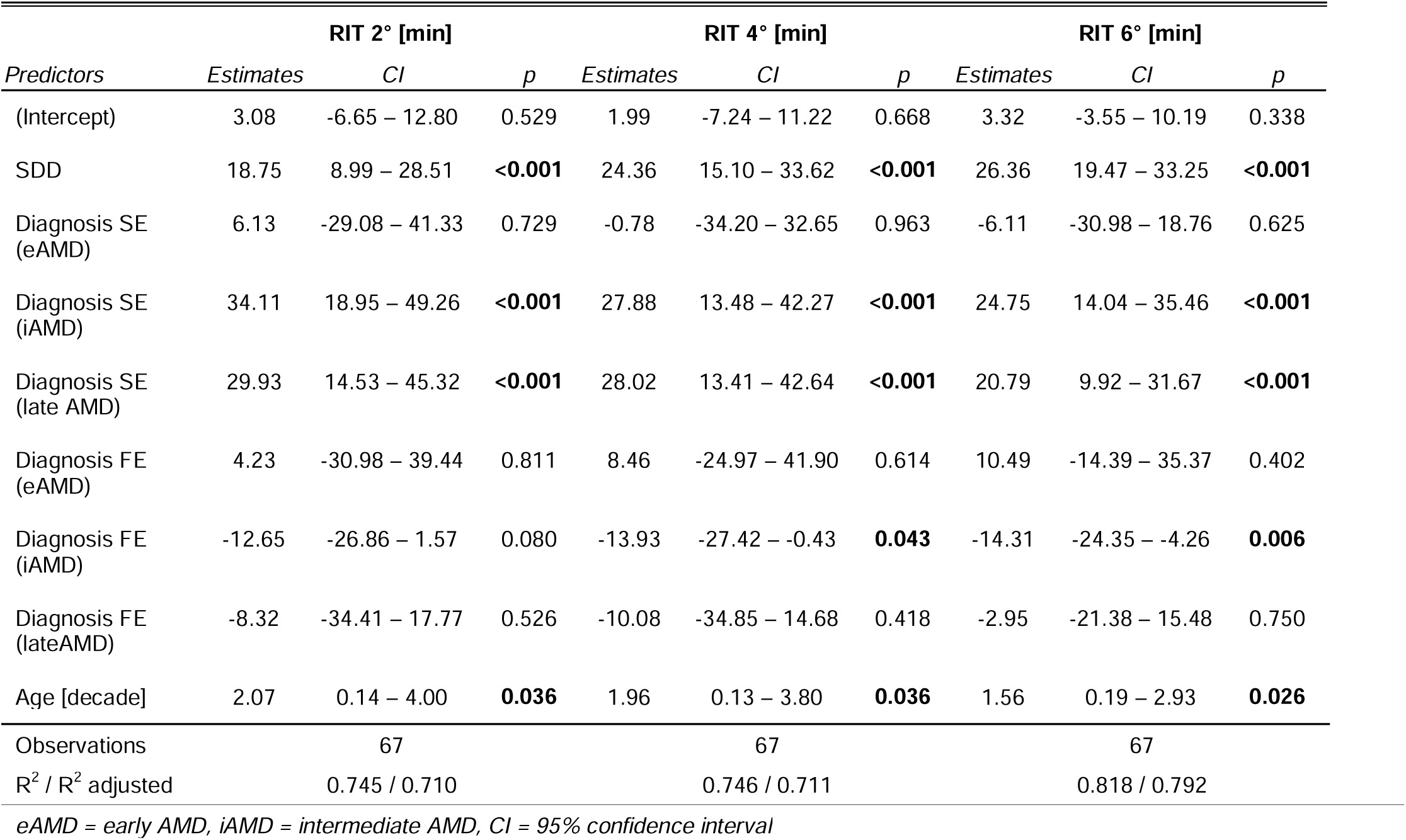
Association between Rod Intercept Time (RIT) and Subretinal Drusenoid Deposits (SDD) as well as AMD-Severity in the Study Eye (SE) and the Fellow-Eye (FE), with Adjustment for Age.

| Predictors | RIT 2° [min] |  |  | RIT 4° [min] |  |  | RIT 6° [min] |  |  |
| --- | --- | --- | --- | --- | --- | --- | --- | --- | --- |
|  | Estimates | CI | p | Estimates | CI | p | Estimates | CI | p |
| (Intercept) | 3.08 | -6.65 – 12.80 | 0.529 | 1.99 | -7.24 – 11.22 | 0.668 | 3.32 | -3.55 – 10.19 | 0.338 |
| SDD | 18.75 | 8.99 – 28.51 | **<0.001** | 24.36 | 15.10 – 33.62 | **<0.001** | 26.36 | 19.47 – 33.25 | **<0.001** |
| Diagnosis SE (eAMD) | 6.13 | -29.08 – 41.33 | 0.729 | -0.78 | -34.20 – 32.65 | 0.963 | -6.11 | -30.98 – 18.76 | 0.625 |
| Diagnosis SE (iAMD) | 34.11 | 18.95 – 49.26 | **<0.001** | 27.88 | 13.48 – 42.27 | **<0.001** | 24.75 | 14.04 – 35.46 | **<0.001** |
| Diagnosis SE (late AMD) | 29.93 | 14.53 – 45.32 | **<0.001** | 28.02 | 13.41 – 42.64 | **<0.001** | 20.79 | 9.92 – 31.67 | **<0.001** |
| Diagnosis FE (eAMD) | 4.23 | -30.98 – 39.44 | 0.811 | 8.46 | -24.97 – 41.90 | 0.614 | 10.49 | -14.39 – 35.37 | 0.402 |
| Diagnosis FE (iAMD) | -12.65 | -26.86 – 1.57 | 0.080 | -13.93 | -27.42 – -0.43 | **0.043** | -14.31 | -24.35 – -4.26 | **0.006** |
| Diagnosis FE (lateAMD) | -8.32 | -34.41 – 17.77 | 0.526 | -10.08 | -34.85 – 14.68 | 0.418 | -2.95 | -21.38 – 15.48 | 0.750 |
| Age [decade] | 2.07 | 0.14 – 4.00 | **0.036** | 1.96 | 0.13 – 3.80 | **0.036** | 1.56 | 0.19 – 2.93 | **0.026** |
| Observations | 67 |  |  | 67 |  |  | 67 |  |  |
| R<sup>2</sup> / R<sup>2</sup> adjusted | 0.745 / 0.710 |  |  | 0.746 / 0.711 |  |  | 0.818 / 0.792 |  |  |

eAMD = early AMD, iAMD = intermediate AMD, CI = 95% confidence interval

## DISCUSSION

We found that fundus-tracked dark adaptometry using small, spatially precise stimuli reliably quantifies rod-recovery kinetics in the parafovea that these dynamic parameters outperform steady-state thresholds. The results indicate that a mechanism-linked, spatially appropriate functional biomarker can enhance early disease staging and may help guide interventions aimed at modifying the degenerative trajectory before atrophy develops.

We used fundus-controlled testing at 2°, 4°, and 6° eccentricities, modeling full dark-adaptation curves to derive RIT, final (rod) threshold, and cone threshold (CT), which revealed that RIT abnormalities were nearly ubiquitous at 2° (89%), declining gradually at 4° (77%) and 6° (74%). Final (rod) threshold abnormalities were less common (51% at 2°, 29% at 6°), and cone threshold abnormalities were rare (17 to 23% across loci). Median RIT at 2° was 60 minutes, but less delayed at 4° and 6°. Jointly, these data establish a clear parafoveal to mid-macula gradient, with the parafoveal locations showing the earliest and most frequent dysfunction. Prior work had hinted that dark adaptation deficits increase toward the fovea, but earlier free-viewing perimetry devices could not test closer than 3°,^18^ 4°,^21,35,36^ or even farther eccentric.^22^ The recent development of fundus-tracked dark adaptometry with Goldmann III–sized stimuli ^37^ now enabled us to test patients in a spatially specific manner and confirm that diagnostic accuracy peaks parafoveally (2°). This locus is consistent with the spatial annulus of peak rod photoreceptor loss in histopathology,^14^ as well as retinal thinning observed in healthy eyes with a genetic susceptibility to AMD in UK Biobank.^38^

Dynamic dysfunction (in terms of RIT delay) consistently outperformed steady-state thresholds in differentiating AMD from controls. Notably, the relationship between RIT delay and final (rod) threshold was nonlinear: typically, steady-state rod dysfunction was mostly confined to locations with RIT delays of >40 minutes. A similar trend was seen for the relationship between RIT and cone thresholds. This supports the longstanding hypothesis that dynamic dysfunction precedes steady-state dysfunction in AMD. However, most earlier studies were limited by testing protocols that terminated before final rod thresholds were measurable and did not provide confirmatory data. Flynn et al. demonstrated that dark adaptation phenotypes could be defined (normal, isolated dynamic delay, or combined dynamic and steady-state impairment) but did not determine a quantitative inflection point.^21^ The longitudinal NIH dark adaptation study reported a RIT >22.8 minutes (i.e., RIT delay of 11 minutes with their settings) for the onset of cone threshold elevation,^39^ which differs considerably from our data. This is likely attributable to the unifocal testing at 5° eccentricity (1.7° diameter stimulus), where RIT delays tend to be less pronounced compared to 2°.

Mechanistically similar relationships have been noted in other Bruch’s membrane diseases: L-ORD,^40^ Sorsby fundus dystrophy,^41,42^ and Pseudoxanthoma elasticum,^43^ in which dynamic dysfunction may occur in isolation and steady-state loss only later, when metabolic disruption of the RPE–photoreceptor interface becomes irreversible.

The five minute full-field bleach, corresponding to approximately 59% rhodopsin bleaching, was intentionally set at a relatively high level to enable the detection of subtle functional effects. This value was chosen slightly below previously reported bleaching levels (76%, 82%).^44,45^ Given the robustness of our findings, future studies using fundus-controlled dark adaptometry may achieve comparable discriminating accuracy between early AMD stages even with lower bleaching intensities, similar to the 30% bleach that has been established previously.^21,46^

Regression analyses revealed that subretinal drusenoid deposits were the strongest independent determinant of delayed RIT, with prolongations of +18.6 to +26.3 minutes across eccentricities. These data extend previous findings linking SDD with impaired rod-mediated recovery.^21,47,48^ Interestingly, these models explained 71 to 79% of total RIT variance, despite of the absence of local features. This again is compatible with previous data suggesting that subretinal drusenoid deposits are associated with scotopic dysfunction in a macula-wide manner instead of localized dysfunction.^17,49–51^

### Limitations

Limitations regarding the cohort selection and cross-sectional design of the study must be considered. AMD was classified per eye (allowing us to evaluate fellow-eye influences); however, this resulted in a cohort slightly more severe than a purely bilateral intermediate AMD population, as six participants (22.9%) had late AMD in the fellow eye. Extrapolating from these cross-sectional study results implies that parafoveal (2°) dark adaptometry might provide a superior ability to detect change-over-time compared to mid-macular testing, but longitudinal studies will be required for confirmation. Likewise, the RIT inflection point for steady-state dysfunction will require longitudinal data from confirmation. Additionally, the steady-state values at the end of dark adaptometry are more closely correlated to dark adaptation and should therefore be compared to pre-bleach steady-state values. Despite the excellent explanatory power of the structure-function model based on global features, inclusion of other features – incl. refined severity scales,^52^ hyperreflective foci,^53^ choriocapillaris flow deficits,^54^ and point by point comparison to retinal layer thicknesses^55,56^ – warrants considerations.

Fundus-tracked dark adaptometry provides a spatially precise assessment of rod-mediated dysfunction in AMD. Parafoveal RIT, particularly at 2°, identifies the earliest and most consistent abnormality, marking the transition from delayed rod recovery to steady-state sensitivity loss. These results suggest that parafoveal dark adaptometry captures disease expression at a stage when the retinoid cycle is impaired, but photoreceptor structure remains largely intact. While further longitudinal data are needed, RIT at 2° eccentricity offers a mechanism-linked, quantifiable functional metric with potential value for monitoring early AMD progression and therapeutic response.

## Data Availability

All data are obtained at the University Hospital of Basel, Switzerland. Further enquiries can be directed to the corresponding author.

