## Supplementary Material for "Parafoveal Dark Adaptation in Early and Intermediate Age-Related Macular Degeneration"

**SUPPLEMENT**

**Supplementary Table S1. Priors for the Bayesian Nonlinear Regression**

| **Parameter** | **Unit** | **Priors for S-MAIA data** |
| --- | --- | --- |
| Initial threshold ($t_{0}$) | logUnits | Normal distribution (µ=2.5, SD=1) |
| Exponential cone recovery time constant (τ) | min | Exponential (λ=0.1) |
| Cone threshold ($c_{t}$) | logUnits | Normal distribution (µ=-0.7, SD=2) |
| Cone-rod break time ($CRB$) | min | Log-normal distribution (µ=4, σ=1) |
| S2 slope ($S_{2}$) | logUnits per min | Normal distribution (µ=-0.24, SD=0.05) |
| Final rod threshold ($t_{f}$) | logUnits | Normal distribution (µ=-2.5, SD=1.5) |

**Supplementary Table S2. Cohort Characteristics within AMD-Patients by Presence of Subretinal Drusenoid Deposits (SDD)**

|  | **SDD Absent (N=25)** | **SDD Present (N=10)** |
| --- | --- | --- |
| **Age [years]** |  |  |
| Median [IQR] | 69.1 [61.5, 75.3] | 80.3 [71.3, 83.0] |
| **Sex** |  |  |
| female | 16 (64%) | 6 (60%) |
| male | 9 (36%) | 4 (40%) |
| **Study eye** |  |  |
| Left | 8 (32%) | 6 (60%) |
| Right | 17 (68%) | 4 (40%) |
| **Study eye visual acuity [logMAR]** |  |  |
| Median [IQR] | -0.07 [-0.07, 0.1] | 0.02 [-0.07, 0.17] |
| **Diagnosis: Study eye** |  |  |
| eAMD | 8 (32%) | 3 (30%) |
| iAMD | 16 (64%) | 4 (40%) |
| late AMD: GA | 1 (4%) | 3 (30%) |
| **Diagnosis: Fellow eye** |  |  |
| eAMD | 8 (32%) | 2 (20%) |
| iAMD | 14 (56%) | 3 (30%) |
| late AMD: GA | 3 (12%) | 3 (30%) |
| Late AMD: nAMD | 0 (0%) | 2 (20%) |

*eAMD = early AMD, iAMD = intermediate AMD, nAMD = neovascular AMD, GA = geographic atrophy*

**Supplementary Table S3. Dark Adaptometry Parameters within AMD Patients by Presence of Subretinal Drusenoid Deposits (SDD)**

|  | **SDD Absent (N=25)** | **SDD Present (N=10)** |
| --- | --- | --- |
| **Cone threshold at 2° [logUnits]** | | |
| Median [IQR] | -0.58 [-0.68, -0.43] | -0.41 [-0.58, -0.36] |
| **Final (rod) threshold at 2° [logUnits]** | | |
| Median [IQR] | -1.65 [-2.05, -0.95] | -0.41 [-0.58, -0.36] |
| **Rod intercept time (RIT) at 2° [min]** | | |
| Median [IQR] | 33.1 [22.7, 60.0] | 60.0 [60.0, 60.0] |
| **Cone threshold at 4° [logUnits]** | | |
| Median [IQR] | -0.57 [-0.66, -0.37] | -0.35 [-0.53, -0.15] |
| **Final (rod) threshold at 4° [logUnits]** | | |
| Median [IQR] | -2.14 [-2.35, -1.81] | -0.54 [-1.30, -0.25] |
| **Rod intercept time (RIT) at 4° [min]** | | |
| Median [IQR] | 24.4 [15.6, 41.8] | 60.0 [60.0, 60.0] |
| **Cone Threshold at 6° [logUnits]** | | |
| Median [IQR] | -0.49 [-0.57, -0.35] | -0.32 [-0.36, -0.30] |
| **Final (rod) threshold at 6° [logUnits]** | | |
| Median [IQR] | -2.33 [-2.45, -2.08] | -0.87 [-1.99, -0.13] |
| **Rod intercept time (RIT) at 6° [min]** | | |
| Median [IQR] | 21.2 [12.9, 38.6] | 60.0 [45.7, 60.0] |

**Supplementary Table S4. Prevalence of Dark-Adaptation Abnormalities by Eccentricity: Evidence of a Parafoveal Gradient in AMD**

Abnormal cone threshold (CT), final rod threshold (FT), and rod intercept time (RIT) were reported as rates outside age-adjusted normal limits.

| **N** | **Position** | **Abnormal CT [rate]** | **Abnormal FT [rate]** | **Abnormal RIT [rate]** |
| --- | --- | --- | --- | --- |
| 35 | 2° | 0.17 | 0.51 | 0.89 |
| 35 | 4° | 0.20 | 0.37 | 0.77 |
| 35 | 6° | 0.23 | 0.29 | 0.74 |

**Supplementary Figure S1. Example of Fundus-controlled Two-color Dark Adaptation at 4°** The study participant was a female 72-year-old with intermediate AMD with a BCVA of -0.07 logMAR and no subepithelial drusenoid deposits. Sky-blue lines represent fitted model curves. Cone-rod break ($CRB$) is shown by the dashed line, rod intercept time (RIT) was defined as the time to detect a criterion stimulus of -1.4 logUnits, S2-slope ($S_{2}$) is represented by the red line, cone threshold ($c_{t}$) and final (rod) threshold ($t_{f}$) are marked on the right.

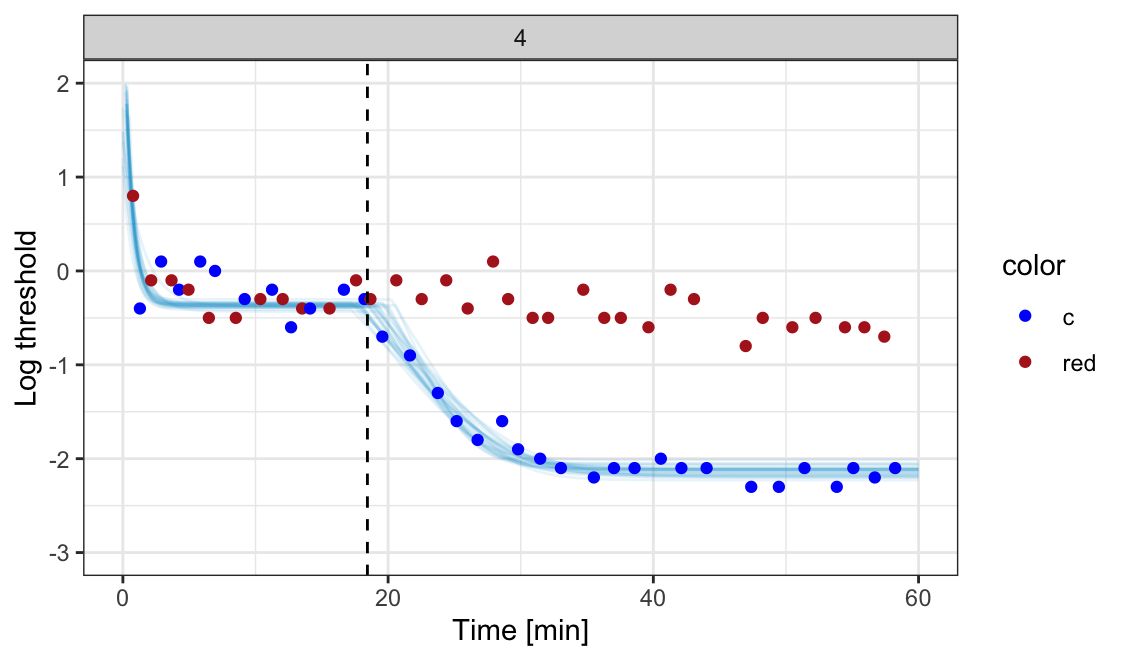

$$c_{t}$$

$$CRB$$

$$t_{f}$$

$$\mathrm{RIT}$$

$$S_{2}$$

**COMMERCIAL RELATIONSHIPS DISCLOSURES:**

Georg Ansari:	BrightFocus Foundation (R), iCare (R), F.Hoffmann-La Roche (R)

Jeannine Oertli:	None (N)

Laura Mächler:	None (N)

Theresa Lipsky:	None (N)

Nicolas Feltgen:	AbbVie (C), Bayer (C), Chiesi (C), Heidelberg Engineering (C), Novartis (C), F.Hoffmann-La Roche (C)

Brett G Jeffrey:	None (N)

Catherine A Cukras:	F. Hoffmann-La Roche (E)

Caroline C W Klaver:	AbbVie (R), Bayer (R), Topcon (R)

Kristina Pfau:	Daiichi Sankyo (C), Bayer (R), Heidelberg Engineering (R), F. Hoffmann-La Roche (R)

Maximilian Pfau:	F. Hoffmann-La Roche (E), Novartis (R), Apellis (C, F), iCare (F)

**FUNDING SOURCES**

This project was supported by the BrightFocus Foundation (M2024009N to Maximilian Pfau).

**DATA AVAILABILITY STATEMENT**

All data are obtained at the University Hospital of Basel, Switzerland. Further enquiries can be directed to the corresponding author.
